# Using FIB-4 score as a screening tool in the assessment of significant liver fibrosis (F2) in patients with transfusion dependent beta thalassaemia: a cross sectional study

**DOI:** 10.1101/2022.01.15.22269369

**Authors:** Padmapani Padeniya, Dileepa Ediriweera, Arjuna De Silva, Madunil Niriella, Anuja Premawardhena

**Affiliations:** Faculty of Medicine, University of Kelaniya, Ragama, Sri Lanka; Hemas Hospital, Wattala, Sri Lanka; Adolescent and Adult Thalassaemia Care Center (University Medical Unit), North Colombo Teaching Hospital, No. 10, Sirima Bandaranayake Mawatha, Kadawatha, Kadawatha, Sri Lanka; Nawaloka Hospital PLC, Colombo, Sri Lanka

**Keywords:** Significant liver fibrosis, Transient elastography, FIB-4 score, Thalassemia, transfusion-dependent anaemia

## Abstract

**Objective:** To evaluated the performance of FIB-4 score as a screening tool to detect significant liver fibrosis (F2) compared to transient elastography (TE), among chronic transfusion-dependent beta-thalassemia (TDT) patients, in a resource-poor setting.

**Design:** A cross-sectional study

**Setting:** Adolescent and Adult Thalassaemia Care Center (University Medical Unit), Kiribathgoda, Sri Lanka.

**Participants:** 45 TDT patients who have undergone more than 100 blood transfusions with elevated serum ferritin >2000ng/mL, were selected for the study. Patients who were serologically positive for hepatitis C antibody were excluded.

**Outcome measures:** TE and FIB-4 score were estimated at the time of recruitment in all participants. Pre-defined cut-off values for F2, extracted from previous studies for TE and FIB-4 score, were compared. A new cut-off value for FIB-4 score was estimated using ROC curve analysis to improve the sensitivity for F2 prediction.

**Results:** Of the selected 45 TDT patients, 22(49%) were males. FIB-4 score showed a significant linear correlation with TE (r= 0.52;*p*< 0.0003). The FIB-4 score was improbable to lead to a false classification of TDT patients to have F2 when the FIB-4 cut-off value was 1.3. On the other hand, it had a very low diagnostic yield in missing almost all (except one) of those who had F2. Using a much-lowered cut-off point of 0.32 for FIB-4, we improved the pick-up rate of F2 to 72%.

**Conclusions:** Regardless of the cut-off point, FIB-4 score cannot be used as a good screening tool to pick-up F2 in patients with TDT, irrespective of their splenectomy status. On the contrary, at 1.3 cut off value though FIB-4 is a very poor detector for F2 fibrosis it will not erroneously diagnose F2 fibrosis in those who don’t have it.

**Article summary:** *Strengths and limitations of this study:*

- There is limited information available on the applicability of FIB-4 score to assess significant liver fibrosis in patients with transfusion-dependent beta-thalassemia (TDT).
- The present study among a Sri Lankan TDT population had their liver fibrosis assessed by FIB-4 score biomarker compared with Transient elastography (TE).
- Small sample size of the study is one of the major limitations.
- Even though the liver biopsy is the gold standard method of assessing liver fibrosis, non-invasive TE was used as the reference standard in our study

## Introduction

Liver disease is the third commonest cause of morbidity and mortality in transfusion-dependent thalassaemia (TDT) patients. Though infections and cardiac failure are the leading causes of death among these patients liver is yet another major target organ susceptible to damage ^1^. Transfusional iron overload and transfusion-transmitted hepatitis infections are the leading causes of hepatic fibrogenesis triggering liver cell dysfunction in these patients ^2 3^. Upon chronic exposure to excess iron, myofibroblasts activate and secrete extracellular matrix protein, predominantly collagen type I and III, assisting in scar tissue formation, causing liver fibrosis. ^4 5^.

Assessing liver fibrosis utilizing liver biopsy is the gold standard method. Owing to its procedure related mortality (<1 in 10,000 cases) and its invasive nature, liver biopsy is not favoured by clinicians or patients ^6 7 8^. Transient elastography (TE) (FibroScan^®^) estimates liver fibrosis/stiffness non-invasively. This technology was first introduced by *Sandrin et al*. in 2003. It is a rapid bedside tool with remarkable reproducibility ^9^. TE is based on measuring the velocity of a mechanical shear wave generated by a transducer placed on the skin. The shear wave velocity is decided by the time the shear wave takes to travel through the liver tissue, and the velocity is then converted to liver stiffness measurement and is expressed in kilopascals (kPa). ^10 7 11 12 13^. Despite being simple, safe, and efficient, routine use of TE is restricted by the cost of the device, primarily in resource-poor settings.

Consequently, the desire to develop clinical scores to detect liver fibrosis using inexpensive point of care tests have always been explored ^8^. Of the several unique biomarkers that have been developed, the FIB-4 score has been widely used. This was initially developed in patients with HIV/HCV coinfection to predict liver fibrosis and has been validated in HCV and non-alcoholic fatty liver disease (NAFLD) patients. It is a simple model based on biochemical parameters of alanine aminotransferase (ALT), aspartate aminotransferase (AST) and platelet count, along with the age of the patient ^14 15^. Though its interpretation is cautious in splenectomised thalassaemia patients due to post splenectomised thrombocytosis, the FIB-4 score is a cost-effective bedside biomarker for liver fibrosis assessment in primary care. However, there is limited information available on the applicability of this score in patients with TDT ^16 17^. Moreover, data regarding diagnostic performance of FIB-4 score and its comparability with TE in TDT patients is scarce.

The objective of this study was to evaluate the performance of the FIB-4 score in assessing liver fibrosis compared to TE and assess if the FIB-4 score could be used as a screening tool to detect significant liver fibrosis (F2), among chronic transfusion-dependent beta-thalassemia patients, in a resource-poor setting.

## Methodology

### Study design

We prospectively followed up a selected cohort of patients with TDT undergoing aggressive chelation therapy over two years to assess the variation of liver fibrosis. This paper is a cross-sectional study on the baseline characteristics of the study participants at the time of recruitment.

### Study setting and the participants

Patients in our cohort were registered at the Adult and Adolescent Thalassaemia Unit, Kiribathgoda, Sri Lanka. Written informed consent was obtained from each study participant/ guardian at the time of recruitment. Ethical clearance was obtained from the Ethics Review Committee (ERC) of the Faculty of Medicine, University of Kelaniya, Sri Lanka. The study was performed following the declaration of Helsinki.

TDT patients who have undergone more than 100 blood transfusions with elevated serum ferritin >2000ng/mL on three consecutive occasions, three months apart, were selected for the study. Patients who were serologically positive for hepatitis C antibody were excluded from the study. At the time of enrolment, blood was taken for the laboratory evaluation of full blood count (FBC), ALT and AST. All the patients underwent TE to quantify liver fibrosis which was carried out by the same operator in all patients.

### Transient elastography

TE was performed as per the manufacturer’s instructions using FibroScan^®^ (Echosens™, Paris, France). Relevant clinical guidelines for the elastography assessment were referenced in accordance with Ferraioli *et al*. (2016). The median value of at least ten valid measurements with >60% success rate (ratio of valid measures to the total number of measures) and an interquartile range of <30% of the median liver stiffness measurement were considered as successful TE scores. A cut-off value for TE scores to estimate significant liver fibrosis (F2) was pre-defined as per the previously published literature. According to Ferraioli *et al*., a study done on patients with beta thalassaemia TE value of 7.0 kPa was considered the cut off value for significant liver fibrosis (F2) ^12^. Therefore, the current study followed the same threshold of 7 kPa to estimate F2 fibrosis in patients with TDT.

### FIB-4 score

FBC, AST and ALT values at the time of recruitment were considered for the FIB-4 score estimation. The age of the patient was calculated according to their last birthday. FIB-4 score was estimated in all patients according to the following published formula; age (years) * AST [U/L] / (platelets [109/ L] * (ALT [U/L])1/2) ^15 18 19^. Though this score was initially developed in HIV/HCV coinfected patients FIB-4 score has been validated in both HCV and NAFLD patients. According to Castera *et al*. (2019), the prespecified cut off value to rule out F2 fibrosis in patients with NAFLD is 1.3 ^20^. Conversely, Sterling *et al. (*2006) reported that a cut-off value of 1.45 with a negative predictive value of 90% was considered the threshold level for predicting F2 fibrosis in patients with HIV/HCV coinfection ^15^. When we recruited the study participants, we excluded transfusion-dependent beta thalassaemia patients who had been infected with the hepatitis C virus.Hence our study followed the threshold level of 1.3, which is similar to NAFLD patients, to rule out significant liver fibrosis.

### Statistical analysis

Data analysis was carried out in two stages. During the first stage, the spearman correlation coefficient was estimated between the FIB-4 and TE scores to find an association between the two measurements in the study group. To assess the FIB-4 score as a screening tool to detect F2 fibrosis with reference to TE findings, cut off values were pre-defined for FIB-4 and TE scores as 1.3 and 7 kPa, respectively. In the entire study population, the sensitivity and specificity of the FIB-4 score as a screening tool to detect significant fibrosis was calculated and tabulated. During the second stage, receiver operating characteristics (ROC) curve analysis was undertaken to identify a new cut off value with good sensitivity for the FIB-4 score.

The distribution of continuous variables was expressed as mean (SD), and categorical variables were presented as frequencies. The *p-*value <0.05 was considered to be statistically significant. All descriptive and analytical statistics were calculated with R programming language version 3.4.2.

## Results

Of the 45-transfusion dependent beta thalassaemia patients we studied, 22 (49%) were males. The mean age of the patients was 18.9 years (SD = 4.8). Thirty (67%) patients have not undergone splenectomy in our study group. Of the 15 (33%) patients who had undergone splenectomy seven (47%) patients had thrombocytosis (defined as >450,000) following post splenectomy. Demographic, biochemical characteristics, FIB-4 and TE scores of the entire group, the un-splenectomised group and the splenectomised group are summarized in table 1. There were no failures recorded in the TE assessment.

**Table 1:**
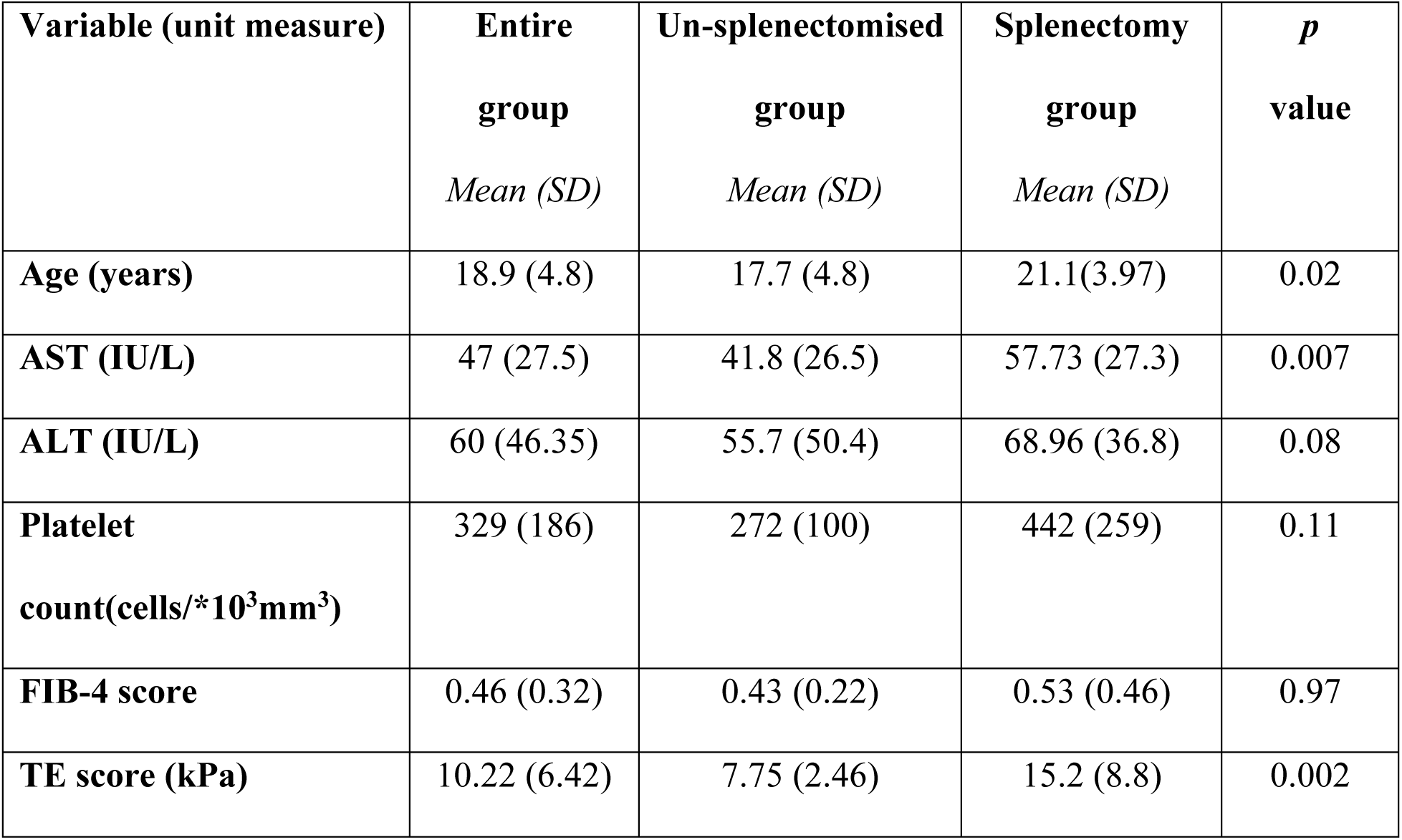
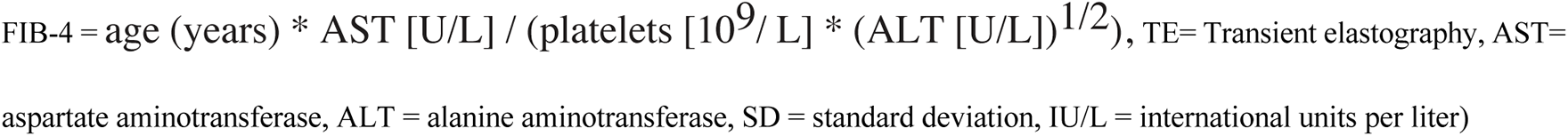
Demographic and biochemical characteristics, FIB- 4 and TE scores of the study participants.

| **Variable (unit measure)** | **Entire group**<br>*Mean (SD)* | **Un-splenectomised group**<br>*Mean (SD)* | **Splenectomy group**<br>*Mean (SD)* | ***p* value** |
| --- | --- | --- | --- | --- |
| **Age (years)** | 18.9 (4.8) | 17.7 (4.8) | 21.1(3.97) | 0.02 |
| **AST (IU/L)** | 47 (27.5) | 41.8 (26.5) | 57.73 (27.3) | 0.007 |
| **ALT (IU/L)** | 60 (46.35) | 55.7 (50.4) | 68.96 (36.8) | 0.08 |
| **Platelet count(cells/\*10<sup>3</sup>mm<sup>3</sup>)** | 329 (186) | 272 (100) | 442 (259) | 0.11 |
| **FIB-4 score** | 0.46 (0.32) | 0.43 (0.22) | 0.53 (0.46) | 0.97 |
| **TE score (kPa)** | 10.22 (6.42) | 7.75 (2.46) | 15.2 (8.8) | 0.002 |

FIB-4 = age (years) \* AST [U/L] / (platelets [ $10^9$ /L] \* (ALT [U/L])<sup>1/2</sup>), TE= Transient elastography, AST= aspartate aminotransferase, ALT = alanine aminotransferase, SD = standard deviation, IU/L = international units per liter)

The FIB-4 score showed a significant linear correlation with TE scores in the entire study group (r = 0.52; *p* < 0.0003).

At the time of recruitment, there were 29 (64%) patients who had significant liver fibrosis as per the TE results. Conversely, the FIB-4 score detected only 1 out of 29 patients with significant liver fibrosis in the study group; hence sensitivity is 3.4% (table 2). In other words, when a pre-defined cut off value of 1.3 was set for FIB4 score, only 1 (1/29) patient was correctly classified as having significant liver fibrosis in our study cohort (true positives). Twenty-eight out of 29 patients (28/29) were misclassified as not having significant liver fibrosis (false negative). Similarly, at the time of recruitment, 16 patients did not have significant liver fibrosis as per the TE assessment (true negatives). Of that group, the FIB-4 score was able to classify 100% as not having significant liver fibrosis; hence specificity is 100%. Not a single patient without significant liver fibrosis was misclassified as having significant liver fibrosis (false positives). At this cut off point, it’s a very poor detector for fibrosis but will not erroneously diagnose fibrosis in those who don’t have it.

**Table 2:**
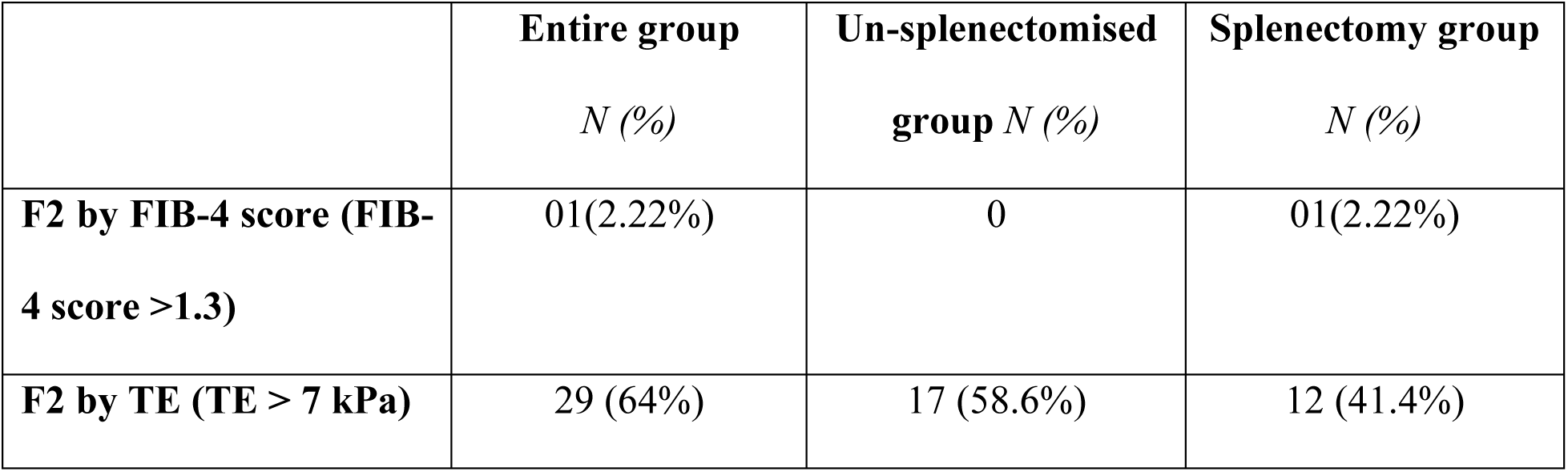
At the time of recruitment, number of patients with significant liver fibrosis (F2) by FIB-4 score and TE assessment.

Table 3 shows the sensitivity, specificity and area under the curve (AUC) for the FIB-4 score with a new cut off value to detect F2 fibrosis compared to the pre-defined cut off value of 1.3.

**Table 3:** ROC curve analysis data in the total study participants in comparison with the previously published FIB-4 cut off value of 1.3.

| Study group | FIB-4 cut-off value |  | AUROC | Sensitivity | Specificity |
| --- | --- | --- | --- | --- | --- |
| Entire group | As per the pre-defined cut off value | 1.3 | - | 3.4% | 100% |
|  | New cut off value from ROC curve analysis | 0.32 | 66% | 72% | 56% |

(AUROC: Area under receiver operating characteristics)

The best new cut off point to detect significant liver fibrosis in this study group is 0.32 (AUC = 66%) (figure 1). With the suggested new cut off value of 0.32, the FIB-4 score was able to detect 21 out of 29 patients (72%) with F2 fibrosis in the study cohort. Hence sensitivity improved from 3.4% to 72 %. Only 8 out of 29 patients (8/29) were misclassified as not having significant liver fibrosis (false negative). As per the new cut off value, nine patients were correctly classified as having F2 fibrosis (true negatives), whereas seven patients were misclassified as not having F2 fibrosis (false positives). Hence the new cut-off value is unable to rule out patients with significant liver fibrosis 100%.

**Figure 1:**
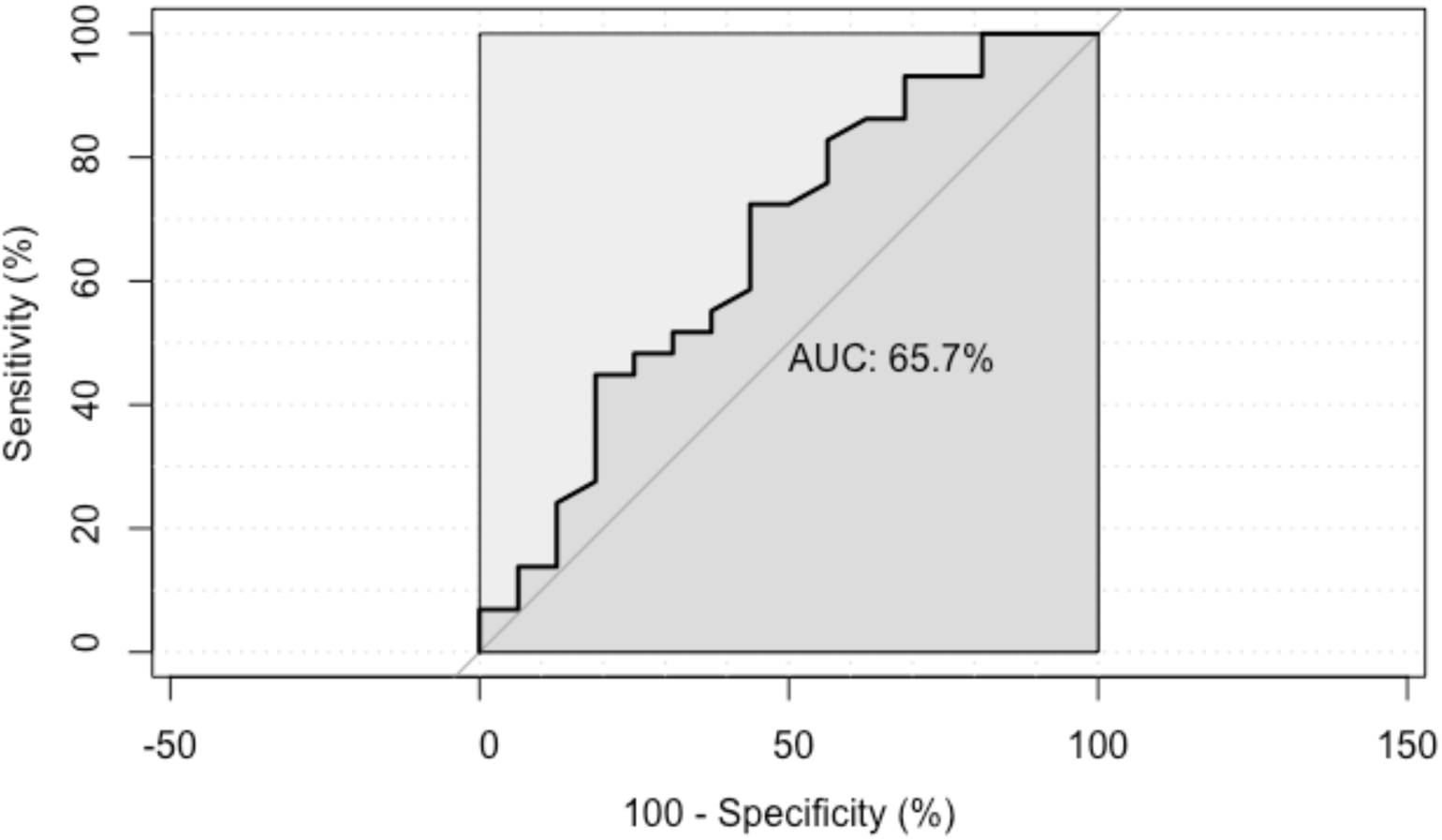
ROC curve analysis of the total study participants (AUC = 66% with the best new cut-off for FIB-4 score = 0.32) (ROC: Receiver operating characteristics, AUC: area under the curve)

## Discussion

Hepatic transient elastography is one of the most useful imaging modalities for liver fibrosis assessment. Though this technology was first validated in patients with chronic viral hepatitis C infection, it has not been validated for use in the thalassaemia population ^21^. Nevertheless, TE has been recommended as a reliable tool in assessing liver fibrosis in patients with transfusion-dependent thalassaemia ^22 23 24 25^ While the FIB-4 score was initially developed and validated in patients with HIV/HCV coinfection to predict liver fibrosis, this score has previously been tested only on a few occasions in patients with transfusion-dependent thalassaemia ^15 17^. Yet as per the WHO recommendation, the FIB-4 score can be applied to assess hepatic fibrosis in resource-poor settings when TE is not feasible ^26^.

Of the 15 patients who had undergone splenectomy in our study cohort, 7 (47%) patients had thrombocytosis following post-splenectomy. Post-operative thrombocytosis is a known complication in beta thalassaemia major patients following splenectomy. Approximately seventy-five per cent of splenectomised patients developing thrombocytosis even reaching values as high as 1,000,000 cells/mm^3^ is known ^23 27^. In our study cohort, there is no statistically significant difference between the mean platelet count of the splenectomised and un-splenectomised patient groups (*p*=0.11). Therefore, in the FIB-4 calculator, though the platelet count is a key component, changes of which would unlikely to affect the final result of this study cohort.

With the ROC curve analysis, we propose a better cut off value of 0.32 with 72% sensitivity and 56% specificity (Table 3; Figure 1) for the FIB-4 score in patients with TDT. Yet the AUROC of 66% does not satisfactorily predict F2 fibrosis in this patient cohort. The findings of this study are consistent with the study done by Hamidieh *et al*. on paediatric patients with transfusion-dependent thalassaemia. The study has concluded that TE was superior to the FIB-4 index in fibrosis assessment, and the best cut off value of 0.699 shows AUROC of 61% in paediatric thalassaemia major patients ^17^. Besides a study done on 76 hepatitis C virus-infected patients with beta thalassaemia, Poustchi et al. (2013) disclosed a cut-off value of 0.25 with an AUROC of 51% to detect significant liver fibrosis ^16^.

The data from the present study suggests that when using TE as the standard (in the absence of liver biopsy) and using the FIB-4 score cut off value of 1.3, the FIB-4 score is improbable to lead to a false classification of thalassaemia patients to have significant fibrosis (F2). On the other hand, it had a very low diagnostic yield in missing almost all (except one) of those who had significant fibrosis. Using a much lowered cut off point for FIB-4 score at 0.32, we were able to improve the pick-up rate of F2 fibrosis using the FIB-4 score to 72%. Yet the new cut off point misclassified seven patients as not having significant liver fibrosis, hence unable to exclude patients with F2 100%.

Regardless of the FIB-4 cut-off point, it is clear that in clinical practice FIB-4 score cannot be used as a good screening tool to pick up significant liver fibrosis in patients with TDT, irrespective of their splenectomy status. On the contrary, at 1.3 cut off value though FIB-4 is a very poor detector of F2 fibrosis it will not erroneously diagnose F2 fibrosis in those who don’t have it.

We acknowledge that the small sample size of our study is one of the significant limitations. A similar study with a larger sample size would be required to justify the findings of this study. Even though the liver biopsy is the gold standard method of assessing liver fibrosis, TE was used as the reference standard in our study. Hence part of the variability of our study findings would have been accounted for this. Yet we believe that TE has been validated in various other clinical conditions. It has shown a very good correlation with the histological gradings of liver fibrosis ^28 29^.

## Data Availability

All data produced in the present study are available upon reasonable request to the authors

## Declarations

### Ethics approval and consent to participate

Ethical clearance was obtained from the Ethics Review Committee (ERC) of the Faculty of Medicine, University of Kelaniya, Sri Lanka and the study was performed in accordance with the declaration of Helsinki.

### Patient Consent for Publication

Not applicable

### Availability of data and material

All data generated or analyzed during this study are included in this published article or uploaded as supplementary information: Not applicable

### Author contributions

Anuja Premawardhena contributed to design and perform the research, data analysis, interpretation and draft the paper and critically revised the paper. Padmapani Padeniya was involving in designing and performing the research, data analysing and drafting the paper. Madunil Niriella was contributing to design the research and critically revised the paper. Dileepa Ediriweera was helping in data analysing and interpretation and drafting the paper. Arjuna De Silva critically revised the paper. All the authors approved the final version of the article.

## Acknowledgements

All the staff members of the Adolescent and Adult Thalassaemia Care Center (University Medical Unit), North Colombo Teaching Hospital, No. 10, Sirima Bandaranayake Mawatha, Kadawatha

